# Diabetes diagnosis, treatment, and control in India: results from a national survey of 1.65 million adults aged 18 years and older, 2019-2021

**DOI:** 10.1101/2023.02.06.23285544

**Authors:** Jithin Sam Varghese, Ranjit Mohan Anjana, Pascal Geldsetzer, Nikkil Sudharsanan, Jennifer Manne-Goehler, Harsha Thirumurthy, Soura Bhattacharya, K.M. Venkat Narayan, Viswanathan Mohan, Nikhil Tandon, Mohammed K. Ali

**Affiliations:** Emory Global Diabetes Research Center of Woodruff Health Sciences Center and Emory University, Atlanta, USA; Hubert Department of Global Health, Rollins School of Public Health, Emory University, Atlanta, USA; Madras Diabetes Research Foundation and Dr Mohan’s Diabetes Specialities Centre, Chennai, India; Division of Primary Care and Population Health, Department of Medicine, Stanford University, Stanford, CA, USA; Chan Zuckerberg Biohub, San Francisco, CA, USA; Professorship of Behavioral Science for Disease Prevention and Health Care, Technical University of Munich, Munich, Germany; Heidelberg Institute of Global Health, Heidelberg University, Germany; Division of Infectious Diseases, Brigham and Women’s Hospital, Harvard Medical School, Boston, USA; Leonard Davis Institute of Health Economics and Perelman School of Medicine, University of Pennsylvania, USA; Lattice Innovations, New Delhi, India; Department of Endocrinology and Metabolism, All India Institute of Medical Sciences, New Delhi, India; Department of Family and Preventive Medicine, School of Medicine, Emory University, Atlanta, USA

**Keywords:** Health system performance, Diabetes management, Low- and middle-income country, Diagnosis, Awareness, Treatment, Control, India, Asian Indians

## Abstract

**Importance:** Diabetes mellitus (DM) is widespread and treatable. Little is known about the diabetes care continuum (diagnosis, treatment, and control) in India, and whether it varies by socio-demographic characteristics and vary at the national, state, and district levels.

**Objective:** To estimate the diabetes care continuum among individuals aged 18-98 years old at national, state, and district-levels, and by socio-demographic group.

**Design:** Cross-sectional, nationally representative survey

**Setting:** 28 states, 8 union territories, and 707 districts of India

**Participants:** 1,895,287 approached in the Fifth National Family Health Survey (NFHS-5), 2019-2021

**Exposures:** District, state, urban vs rural residence, age (18-39, 40-64, ≥65 years), sex, household wealth quintile

**Main Outcomes and Measures:** Diabetes was defined by self-report or high capillary blood glucose (≥126mg/dL [fasting] or ≥220mg/dL [non-fasting]). Of those with diabetes, we estimated proportions that were diagnosed (self-reported). Among those diagnosed, we reported the proportions treated (self-reported medication use) and proportion controlled (blood glucose <126 [fasting] or ≤180 mg/dL [non-fasting; corresponding to HbA1c≤8%]). We benchmarked findings against the World Health Organization’s Global Diabetes Compact Targets (80% diagnosis, 80% control among those diagnosed). We partitioned the variance in indicators between state and district levels using variance partition coefficients (VPC).

**Results:** Among 1,651,176 adult respondents (52.6% female; mean age: 41.6 years) with blood glucose measures, the proportion with diabetes was 6.5% (95%CI: 6.4, 6.6). Among adults with diabetes, 74.2% [73.3, 75.0] were diagnosed. Among those diagnosed, 59.4% [58.1, 60.6] reported taking medication and 65.5% [64.5, 66.4] achieved control. Diagnosis and treatment were higher in urban areas, older age groups, and wealthier households. Of the 707 districts, 34.8% districts met diagnosis target, while 10.7% districts met the control target among those diagnosed. Most of the variability in diabetes diagnosis (VPC:69.9%), treatment (VPC:51.8%), and control (VPC:66.8%) were between districts in a state, and not between states.

**Conclusions and Relevance:** Nationally, the diabetes care continuum masks considerable state- and district-level variation, as well as age- and rural-urban disparities. Surveillance at the district-level can guide state health administrators to prioritize interventions and monitor achievement of global targets.

**KEY POINTS:** *Question:* How does the diabetes care continuum (diagnosis, treatment, and control) vary by state, district, and sociodemographic groups in India?

*Findings:* Nationally, among 1.65 million respondents in the National Family Health Survey of 2019-2021, 74.2% were diagnosed. Among those diagnosed, 59.4% reported taking medication and 65.5% achieved control. Most of the variability in care continuum was within-state, between districts (% variance explained: 51.8-69.9) and not between-states. Higher diagnosis and treatment, but lower control was observed in urban compared to rural areas and older compared to younger and middle age groups.

*Meaning:* Considerable differences between states, between districts in a state, for rural adults, and by age imply the need for targeted, decentralized solutions to improve the diabetes care continuum in India.

## Introduction

India is now the most populous country in the world and diabetes affects 74 million residents, representing 14% of the global burden of disease.^1^ Strong evidence for interventions to improve diagnosis, treatment, and control exist to mitigate the complications of diabetes.^2, 3^ Several high-income nations have implemented quality of care programs for diabetes, and achieved reductions in population level rates of diabetes complications.^4, 5^ However, the extent of implementation of these programs in low- and middle-income nations is not clear.

India launched the National Programme for Prevention and Control of Cancer, Diabetes, Cardiovascular Disease and Stroke (NPCDCS), under the National Health Mission in 2010, to prevent and control major non-communicable diseases.^6^ Under the NPCDCS, the National Multi-sectoral Action Plan (NMAP) for Prevention and Control of Common NCDs (2017 to 2022) outlines four priority areas: governance, prevention and promotion, health care and surveillance and monitoring.^7^ In parallel, in May 2021, the World Health Organization (WHO) member states, including India, as part of the World Health Assembly and Global Diabetes Compact, resolved to meet five targets including at least 80% of persons with diabetes should be diagnosed (“diagnosed diabetes”), and 80% of those with diagnosed diabetes have HbA1c levels below 8.0% (“controlled diabetes”).^8^

Monitoring of diabetes quality of care nationally, at subnational levels, and by socio-demographic groups would allow national and state policymakers working under the umbrella of NMAP and NPCDCS to monitor progress and thereby identify priorities for implementing appropriate interventions. While previous studies have estimated the quality of diabetes care, estimates are reported at higher administrative levels (national and state) or among younger adults, impeding decentralized prioritization.^9–13^ Estimates at lower administrative levels and by socio-demographic groups allow targeted solutions to improve the diabetes care continuum.

Using recent nationally representative data, we characterized the diabetes care continuum (diagnosis, treatment, and control) for India, by state, by district, and for each level, by socio-demographic subgroups (sex, urban/rural, age, attained schooling, household wealth).^14–16^ We developed a dashboard for easy visualization of these data at the district level. These data can serve as a basis for driving improvements in diabetes quality of care for India, and will be a valuable resource as the country implements programs and policies toward attaining targets for diagnosis and control recommended by the NMAP and WHO Diabetes Compact.

## Methods

### Study Population

We used data from the National Family Health Survey-5 (NFHS-5), a nationally representative survey conducted in two phases from June 2019 to March 2020, and from November 2020 to April 2021 in 707 districts from 28 states and 8 union territories of India.^17^ NFHS-5 is powered to provide estimates at the district-level.

NFHS-5 used a multi-stage stratified sample where primary sampling units (PSUs) from urban (census enumeration blocks) and rural (villages) strata of each district were sampled at the first stage and households within PSUs were randomly sampled from a list of eligible households at the second stage. NFHS-5 collected data on diabetes status from 612,910 households where eligible participants resided (women: 15-49 years, men: 15-54 years) and were approached.^18^ Household and individual characteristics were collected using standardized instruments.

The survey additionally collected data on blood glucose among all adults (18 years and older) who were living in the same household as eligible participants (pregnant or non-pregnant women 15-49 years and men 15-54 years). However, information such as body mass index or waist circumference were not available for these participants. The overall sample consisted of 1,895,287 adults aged 18-98 years.

We restricted our analytic sample to those who either self-reported having diabetes or who had a valid measurement of blood glucose (**Supplementary Figure 1**). We used district-level boundaries from the NFHS-5 district sampling frame (n = 707).

### Data collection and definitions

#### Diabetes Mellitus

NFHS-5 measured random blood glucose using Accu-Chek Performa glucometers to quantify whole blood glucose from capillary blood.^18, 19^ As reported previously,^20–22^ a conversion factor is used to reliably estimate plasma glucose concentrations (mg/dL).^23^

Diabetes was defined as either self-reported (in response to the question: “Before this survey, were you ever told you had high blood glucose by a doctor, nurse or health practitioner on two or more occasions?”) or based on high blood glucose measurement (≥126 mg/dL if fasting [not eating or drinking anything except water for more than 8 hours] or ≥220 mg/dL if non-fasted). ^22, 24^

#### Diabetes Care Continuum – Diagnosis, Treatment, and Control

We used the following metrics to map the diabetes care continuum: prevalence of diabetes, proportion diagnosed, and among those diagnosed, the proportion treated, and proportion controlled. We defined diagnosed diabetes as proportions of adults with diabetes who reported being diagnosed prior to the survey. Further, among those with diagnosed diabetes, we identified the proportion treated (those self-reporting medication use) and controlled diabetes (those achieving <126mg/dL [fasting] or ≤180mg/dL [not fasting; corresponding to HbA1c≤8%]) per national guidelines for management of Type 2 Diabetes.^24^ We present state-level estimates and the district-level estimates in an interactive online dashboard that accompanies this work. We summarize the definitions in **Supplementary Table 1**.

#### Socio-demographic variables

We used the household wealth index computed as the first principal component from survey responses regarding possession of assets and quality of housing, separately for urban and rural areas, as provided by Demographic and Health Surveys.^25^ We used the following household covariates: rural residence (versus urban) and regional wealth quintile (urban and rural) from the household wealth index as provided by NFHS. We used the following individual strata in our analysis: sex (male or female), age (18-39, 40-64, ≥65 years), and schooling (none or missing, primary [up to 4^th^ class], secondary [up to 10^th^ class], post-secondary).

### Statistical Analysis

We report survey-weighted estimates accounting for the complex survey design and 95% cluster-robust confidence intervals.^17^. We describe the individual and household characteristics of the analytic sample after stratifying by residence (urban or rural) and by sex. We also compared the analytic sample with the excluded (those without data on diabetes status).

Care continuum performance indicators were reported for the total sample for urban and rural areas, as well as stratified by sex, age category, schooling and regional wealth quintile. We reported age-standardized estimates of the performance indicators for different strata at the national-level. We performed age-standardization to the distribution of the total sample since different strata of schooling and wealth have different age distributions. We also report weighted estimates at state-level and district-level that were not age-standardized and relevant for local decision making in this manuscript.

To illustrate the variability between- and within-states (between districts), we present examples from high burden states, namely Kerala and Karnataka. To quantify this variability, we partitioned the variance in prevalence attributable at the district-level using variance partition coefficients from linear mixed models with state-level intercepts.

To further aid policy and priority decision-making, we developed a dashboard to visually depict the disparities in care continuum using Shiny by RStudio. The interactive dashboard can be accessed at: https://egdrc-precision-medicine.shinyapps.io/diabetes_cascade/. We displayed disparities, both crude and age-standardized, by sex and region (Total/Urban/Rural) at the state-level on the “Overview” tab. We compared districts within each state on the “District Disparities” tab. We displayed disparities across socio-demographic characteristics at the state-level on the “Socio-demographic Disparities” tab. All analyses were carried out using R 4.2.0 using srvyr 1.1.1.

## Results

The analytic sample consisted of 1,651,176 adults (men: 783,280; 47.4%, and non-pregnant women: 867,896; 52.6%), representing a response rate of 87.1%. Compared to those who were part of the analytic sample in urban areas (n = 406,463), those in the excluded sample (n=77,941) without data on diabetes were more likely to belong to higher wealth quintiles, and received post-secondary education (**Supplementary Table 2**). In rural areas, the analytic sample (n = 1,244,713) was similar to the excluded sample (n = 166,180). More than half of the analytic sample were under 40 years of age (mean age: 41.6 [41.5, 41.6]), and almost 90% were aged 18-64 years (**Table 1**). Almost two-thirds of the analytic sample lived in rural areas. Less than 1.5% of the analytic sample (women: 1.2% [1.2, 1.3], men: 1.4% [1.3, 1.4]) reported fasting for at least 8 hours before blood glucose measurement.

**Table 1.** Characteristics of participants in analytic sample for estimating care cascade of diabetes in India, n = 1,651,176

|  | **Total** |  | **Urban** |  | **Rural** |  |
| --- | --- | --- | --- | --- | --- | --- |
|  | **Women**<br>(n = 867,896) | **Men**<br>(n = 783,280) | **Women**<br>(n = 212,770) | **Men**<br>(n = 193,693) | **Women**<br>(n = 655,126) | **Men**<br>(n = 589,587) |
| **Age category** |  |  |  |  |  |  |
| 18-39 | 50.8<br>(50.6, 50.9) | 49.7<br>(49.5, 49.9) | 50.1<br>(49.7, 50.5) | 50.2<br>(49.9, 50.6) | 51.1<br>(50.9, 51.3) | 49.4<br>(49.2, 49.7) |
| 40-65 | 39.1<br>(39, 39.2) | 38.4<br>(38.3, 38.6) | 40<br>(39.7, 40.3) | 38.9<br>(38.5, 39.2) | 38.7<br>(38.5, 38.8) | 38.2<br>(38, 38.4) |
| 65 and above | 10.1<br>(10, 10.2) | 11.9<br>(11.8, 12) | 9.9<br>(9.6, 10.1) | 10.9<br>(10.6, 11.1) | 10.3<br>(10.2, 10.4) | 12.4<br>(12.2, 12.5) |
| **Schooling** |  |  |  |  |  |  |
| None | 36.7<br>(36.4, 37) | 17.3<br>(17.1, 17.5) | 22<br>(21.5, 22.5) | 9.4<br>(9.1, 9.7) | 43.5<br>(43.2, 43.8) | 21.1<br>(20.8, 21.3) |
| Primary (up to 4<sup>th</sup> class) | 13.8<br>(13.7, 13.9) | 15<br>(14.8, 15.2) | 12.5<br>(12.2, 12.7) | 11.3<br>(11, 11.6) | 14.4<br>(14.3, 14.6) | 16.8<br>(16.6, 17) |
| Secondary (5<sup>th</sup> to 10<sup>th</sup> class) | 37<br>(36.8, 37.2) | 49.6<br>(49.4, 49.9) | 43.6<br>(43.2, 44) | 50.8<br>(50.4, 51.3) | 33.9<br>(33.7, 34.1) | 49<br>(48.8, 49.3) |
| Post-secondary (11<sup>th</sup> class and above) | 12.5<br>(12.3, 12.7) | 18<br>(17.7, 18.4) | 22<br>(21.5, 22.5) | 28.4<br>(27.8, 29.1) | 8.1<br>(8, 8.3) | 13.1<br>(12.8, 13.5) |
| **Caste of head of household** |  |  |  |  |  |  |
| General or unspecified | 27<br>(26.6, 27.4) | 27.2<br>(26.8, 27.7) | 35.1<br>(34.2, 36) | 35<br>(34.1, 36) | 23.3<br>(22.8, 23.7) | 23.5<br>(23, 24) |
| Other Backward Castes | 42.1<br>(41.7, 42.5) | 41.7<br>(41.3, 42.1) | 41.8<br>(40.9, 42.7) | 41.9<br>(41, 42.8) | 42.2<br>(41.7, 42.7) | 41.6<br>(41.2, 42.1) |
| Scheduled Caste | 21.6<br>(21.2, 21.9) | 21.4<br>(21.1, 21.8) | 19.1<br>(18.3, 19.8) | 19<br>(18.2, 19.7) | 22.7<br>(22.3, 23.1) | 22.6<br>(22.2, 23) |
| Scheduled Tribe | 9.3<br>(9.1, 9.6) | 9.6<br>(9.4, 9.9) | 4.1<br>(3.8, 4.4) | 4.1<br>(3.8, 4.4) | 11.8<br>(11.4, 12.1) | 12.2<br>(11.9, 12.6) |
| **Religion** |  |  |  |  |  |  |
| Hindu | 82.4<br>(82, 82.8) | 82.7<br>(82.3, 83.2) | 78.4<br>(77.5, 79.3) | 79<br>(78.1, 79.9) | 84.3<br>(83.9, 84.7) | 84.5<br>(84.1, 85) |
| Muslim | 12.1<br>(11.7, 12.5) | 11.9<br>(11.4, 12.3) | 15.5<br>(14.6, 16.4) | 15.2<br>(14.3, 16.1) | 10.5<br>(10.1, 10.9) | 10.2<br>(9.8, 10.7) |
| Other | 5.5<br>(5.3, 5.7) | 5.4<br>(5.2, 5.6) | 6.1<br>(5.7, 6.5) | 5.8<br>(5.4, 6.1) | 5.2<br>(5, 5.4) | 5.2<br>(5, 5.4) |
| **Household wealth quintile**<br>**(by residence)** |  |  |  |  |  |  |
| Lowest | 18.6<br>(18.3, 18.9) | 18<br>(17.7, 18.3) | 19.4<br>(18.6, 20.1) | 19.4<br>(18.7, 20.1) | 18.2<br>(17.9, 18.5) | 17.3<br>(17, 17.6) |
| Low | 19.6<br>(19.3, 19.8) | 19.4<br>(19.1, 19.6) | 20.1<br>(19.6, 20.6) | 20.2<br>(19.7, 20.7) | 19.3<br>(19.1, 19.5) | 18.9<br>(18.7, 19.2) |
| Medium | 20.3<br>(20.1, 20.5) | 20.4<br>(20.2, 20.6) | 20.4<br>(19.9, 20.8) | 20.4<br>(19.9, 20.8) | 20.3<br>(20.1, 20.5) | 20.5<br>(20.2, 20.7) |
| High | 20.7<br>(20.4, 20.9) | 21.2<br>(20.9, 21.4) | 20.3<br>(19.8, 20.8) | 20.3<br>(19.8, 20.8) | 20.9<br>(20.6, 21.1) | 21.6<br>(21.2, 21.9) |
| Highest | 20.8<br>(20.5, 21.2) | 21.1<br>(20.8, 21.4) | 19.9<br>(19.2, 20.6) | 19.7<br>(19, 20.4) | 21.3<br>(20.9, 21.7) | 21.7<br>(21.4, 22.1) |
| **Blood glucose**<br>**measurement** |  |  |  |  |  |  |
| Fasting for at least 8 hours | 1.2<br>(1.2, 1.3) | 1.4<br>(1.3, 1.4) | 1.1<br>(1, 1.2) | 1.4<br>(1.3, 1.5) | 1.3<br>(1.3, 1.4) | 1.3<br>(1.3, 1.4) |
| **Diabetes** |  |  |  |  |  |  |
| Self-reported or high blood<br>glucose | 6.1<br>(6, 6.2) | 6.2<br>(6.1, 6.3) | 8.5<br>(8.3, 8.8) | 8.3<br>(8, 8.6) | 4.9<br>(4.8, 5) | 5.2<br>(5.1, 5.3) |
| Self-reported | 4.8<br>(4.7, 4.9) | 4.7<br>(4.6, 4.8) | 7<br>(6.8, 7.2) | 6.6<br>(6.4, 6.9) | 3.8<br>(3.6, 3.9) | 3.8<br>(3.7, 4) |

All values are percentages (95% confidence intervals) accounting for survey design. Estimates are not age-standardized.

### National-level care continuum

The age-standardized proportion [95%CI] with diabetes nationally was 6.5% [95%CI: 6.4, 6.6], and was higher in urban areas (9.7% [9.4, 9.9]) relative to rural areas (4.9% [4.8, 5.0]). The proportion was higher among men (7.2% [7.1, 7.3]) relative to women (5.8% [5.7, 5.9]), and was greater with older age, attained schooling, and household wealth. Similar trends were observed in both urban and rural areas.

Among those with diabetes, 74.2% [73.3, 75.0] reported being diagnosed (**Table 2**). Of those with diagnosed diabetes, 59.4% [58.1, 60.6], and 65.5% [64.5, 66.4], respectively, reported taking medication and had controlled blood glucose. The proportion of those with diagnosed diabetes was higher in urban areas (77.2% [76.0, 78.4]), compared to rural areas (72.2% [71.1, 73.3]). Nationally, and in both urban and rural areas, compared to their respective counterparts, diagnosis was higher among women (**Figure 1**), older age groups, those that attained higher schooling, and those with greater household wealth (**Table 2**).

**Figure 1.**
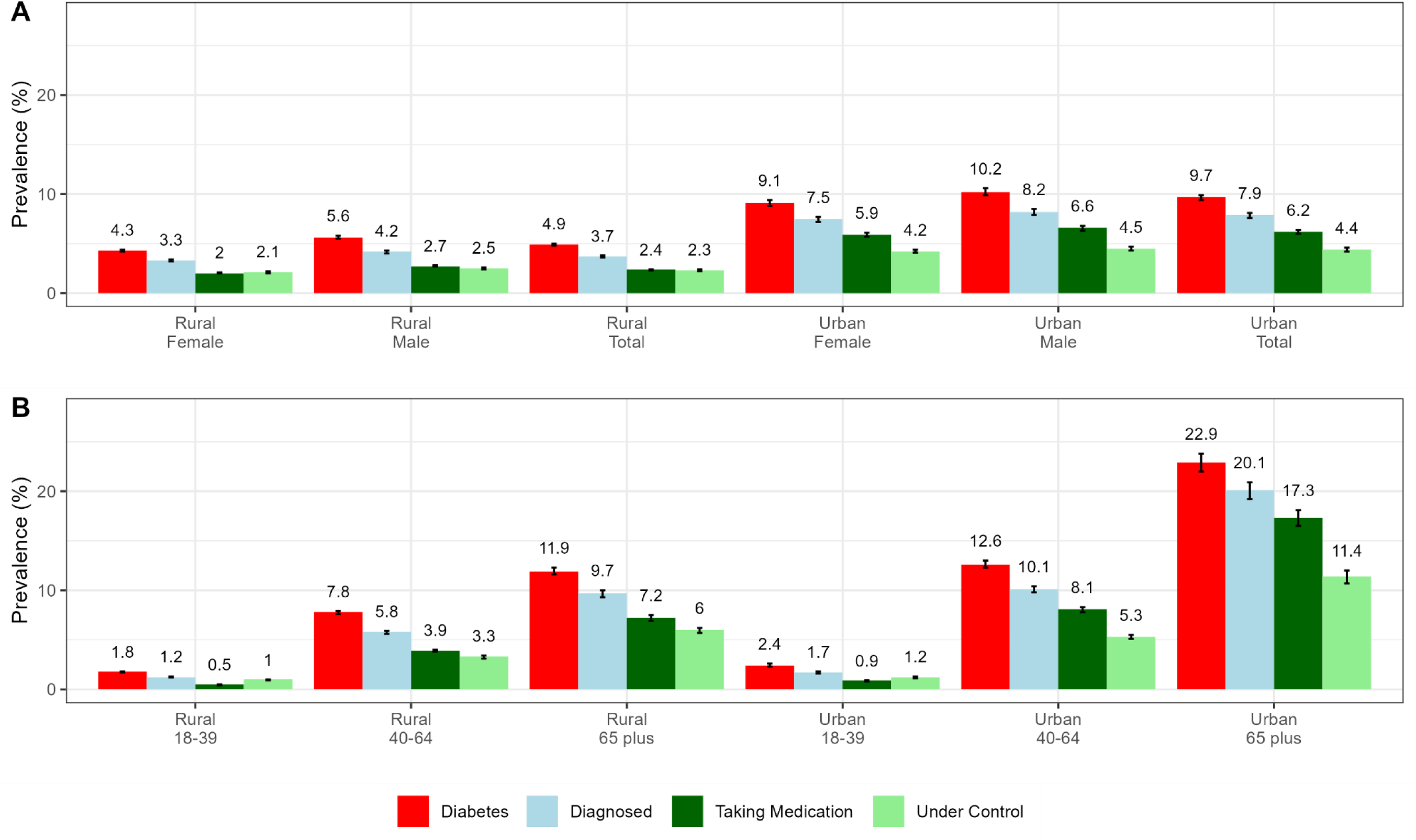
National-level care cascade in analytic sample by urban and rural residence, n = 1,651,176. All values are percentages (error bars: 95% confidence intervals) in total population. Proportions of populations in these categories is presented in **Table 1**. A: Age-standardized and sex-stratified estimates of care cascade; B: Age-stratified estimates of care cascade

**Table 2.** Socio-demographic variations in care cascade in India, n = 1,651,176

|  | **Total** |  |  |  | **Urban** |  |  |  | **Rural** |  |  |  |
| --- | --- | --- | --- | --- | --- | --- | --- | --- | --- | --- | --- | --- |
|  | **Diabetes**<br>**(%)** | **Diagnosed**<br>**a (%)** | **Treated**<br>**b (%)** | **Controlled**<br>**b (%)** | **Diabetes**<br>**(%)** | **Diagnosed**<br>**a (%)** | **Treated**<br>**b (%)** | **Controlled**<br>**b (%)** | **Diabetes**<br>**(%)** | **Diagnosed**<br>**a (%)** | **Treated**<br>**b (%)** | **Controlled**<br>**b (%)** |
| **Total** | 6.5<br>(6.4, 6.6) | 74.2<br>(73.3, 75) | 59.4<br>(58.1, 60.6) | 65.5<br>(64.5, 66.4) | 9.7<br>(9.4, 9.9) | 77.2<br>(76, 78.4) | 69.4<br>(67.4, 71.5) | 59.6<br>(58.1, 61) | 4.9<br>(4.8, 5) | 72.2<br>(71.1, 73.3) | 52.7<br>(51, 54.3) | 69.4<br>(68.1, 70.6) |
| **Sex** |  |  |  |  |  |  |  |  |  |  |  |  |
| Women | 5.8<br>(5.7, 5.9) | 75.9<br>(74.8, 76.9) | 57<br>(55.5, 58.6) | 67.6<br>(66.4, 68.8) | 9.1<br>(8.8, 9.4) | 78.6<br>(77, 80.2) | 68<br>(65.6, 70.5) | 60.6<br>(58.7, 62.4) | 4.3<br>(4.2, 4.4) | 74.2<br>(72.7, 75.6) | 50<br>(47.9, 52) | 72.1<br>(70.5, 73.7) |
| Men | 7.2<br>(7.1, 7.3) | 72<br>(71, 73.1) | 62.5<br>(61.1, 63.8) | 62.6<br>(61.4, 63.8) | 10.2<br>(9.9, 10.6) | 75.6<br>(74.1, 77.1) | 71.2<br>(69, 73.3) | 58.3<br>(56.4, 60.2) | 5.6<br>(5.5, 5.8) | 69.7<br>(68.3, 71.2) | 56.4<br>(54.6, 58.3) | 65.6<br>(64, 67.2) |
| **Age**<br>**category** |  |  |  |  |  |  |  |  |  |  |  |  |
| 18-39 | 1.9<br>(1.9, 2) | 70<br>(68.5, 71.4) | 43.5<br>(41.5, 45.4) | 75.2<br>(73.6, 76.8) | 2.4<br>(2.3, 2.6) | 70.5<br>(67.9, 73) | 51.2<br>(47.6, 54.8) | 69.2<br>(66.3, 72.1) | 1.8<br>(1.7, 1.8) | 69.7<br>(67.9, 71.5) | 39.9<br>(37.5, 42.2) | 78<br>(76.1, 79.9) |
| 40-65 | 9.6<br>(9.4, 9.8) | 76.9<br>(76.3, 77.5) | 74.2<br>(73.2, 75.1) | 54.4<br>(53.6, 55.3) | 12.6<br>(12.3, 13) | 80.3<br>(79.4, 81.2) | 80.3<br>(78.9, 81.7) | 51.9<br>(50.6, 53.2) | 7.8<br>(7.6, 7.9) | 74.1<br>(73.3, 74.8) | 68.6<br>(67.4, 69.9) | 56.7<br>(55.7, 57.8) |
| 65 and<br>above | 16.3<br>(15.9, 16.7) | 83.8<br>(83.1, 84.5) | 79.6<br>(78.6, 80.6) | 60<br>(58.9, 61.1) | 22.9<br>(22, 23.8) | 87.4<br>(86.3, 88.5) | 85.4<br>(84, 86.8) | 57.6<br>(55.9, 59.3) | 11.9<br>(11.6, 12.3) | 80.3<br>(79.4, 81.3) | 73.6<br>(72.2, 75) | 62.4<br>(61.1, 63.7) |
| **Schooling** |  |  |  |  |  |  |  |  |  |  |  |  |
| None | 4.5<br>(4.4, 4.6) | 69.9<br>(68.1, 71.7) | 58.1<br>(55.8, 60.4) | 66.5<br>(64.5, 68.4) | 7.4<br>(7, 7.7) | 72.8<br>(69.7, 76) | 68.1<br>(63.7, 72.4) | 57.8<br>(54.2, 61.4) | 3.9<br>(3.8, 4) | 68.9<br>(66.7, 71.1) | 54.4<br>(51.6, 57.2) | 69.7<br>(67.3, 72) |
| Primary<br>(up to 4<sup>th</sup><br>class) | 6.2<br>(6.1, 6.4) | 72.8<br>(70.9, 74.8) | 59.5<br>(57.1, 62) | 64.4<br>(62.2, 66.7) | 9.8<br>(9.4, 10.3) | 75.1<br>(71.6, 78.6) | 70.1<br>(66.2, 74) | 57<br>(53.4, 60.7) | 5<br>(4.8, 5.2) | 71.6<br>(69.3, 74) | 54.1<br>(50.8, 57.4) | 68.2<br>(65.4, 71.1) |
| Secondary<br>(5<sup>th</sup> to 10<sup>th</sup><br>class) | 7.2<br>(7, 7.3) | 75.8<br>(74.9, 76.7) | 60.5<br>(59.1, 61.9) | 64<br>(62.9, 65.2) | 10.1<br>(9.8, 10.5) | 78.1<br>(76.7, 79.5) | 70.7<br>(68.5, 72.9) | 58.3<br>(56.5, 60.2) | 5.5<br>(5.3, 5.6) | 74<br>(72.8, 75.3) | 52.5<br>(50.6, 54.3) | 68.5<br>(67, 69.9) |
| Post-secondary<br>(11<sup>th</sup> class<br>and above) | 8.5<br>(8.2, 8.8) | 79.7<br>(78.4, 81.1) | 58.1<br>(56.1, 60.1) | 69.4<br>(67.8, 71) | 10<br>(9.5, 10.5) | 80.9<br>(79.1, 82.6) | 67.1<br>(64.3, 69.9) | 65.8<br>(63.6, 68.1) | 6.3<br>(6, 6.6) | 78<br>(75.9, 80.2) | 44.5<br>(41.5, 47.4) | 74.8<br>(72.6, 76.9) |
| **Household**<br>**wealth**<br>**quintile** |  |  |  |  |  |  |  |  |  |  |  |  |
| Lowest | 4<br>(3.8, 4.2) | 67.7<br>(65.6, 69.7) | 49.2<br>(46.5, 52) | 73.2<br>(71.4, 75) | 6.3<br>(5.9, 6.7) | 69.7<br>(66.5, 72.9) | 57.5<br>(52.7, 62.3) | 66.5<br>(63.6, 69.5) | 2.8<br>(2.6, 2.9) | 65.8<br>(63.1, 68.4) | 41.4<br>(38, 44.8) | 79.6<br>(77.2, 82) |
| Low | 5.3<br>(5.1, 5.5) | 70.8<br>(69, 72.5) | 55.9<br>(53.8, 58.1) | 67.7<br>(65.8, 69.5) | 8.9<br>(8.5, 9.3) | 72.2<br>(69.8, 74.7) | 67.4<br>(64.1, 70.6) | 58.6<br>(55.7, 61.5) | 3.5<br>(3.3, 3.6) | 69.4<br>(67.1, 71.8) | 45.7<br>(42.5, 48.8) | 75.8<br>(73.3, 78.3) |
| Medium | 6.4<br>(6.2, 6.6) | 74.7<br>(73.2, 76.1) | 58.7<br>(56.6, 60.7) | 65.9<br>(64.1, 67.7) | 10.2<br>(9.7, 10.7) | 78.2<br>(76.2, 80.1) | 72.5<br>(69.6, 75.4) | 57.2<br>(54.3, 60.1) | 4.5<br>(4.3, 4.6) | 72<br>(70, 74.1) | 47.5<br>(44.8, 50.3) | 72.8<br>(70.6, 75.1) |
| High | 7.5<br>(7.4, 7.7) | 74.6<br>(73, 76.3) | 61.6<br>(59.5, 63.6) | 65<br>(63.3, 66.7) | 11.1<br>(10.6, 11.6) | 80.1<br>(77.8, 82.4) | 70.8<br>(67.7, 74) | 59.8<br>(57.1, 62.5) | 5.8<br>(5.7, 6) | 71.4<br>(69.2, 73.6) | 55.6<br>(52.9, 58.4) | 68.2<br>(66.1, 70.4) |
| Highest | 8.8<br>(8.6, 9) | 77.5<br>(76.1, 79) | 62.7<br>(60.7, 64.7) | 62.3<br>(60.5, 64) | 11.6<br>(11.1, 12.1) | 81.6<br>(79.2, 84) | 72.4<br>(69.2, 75.6) | 58.8<br>(56.1, 61.5) | 7.6<br>(7.4, 7.8) | 75.7<br>(73.8, 77.6) | 58.2<br>(55.6, 60.8) | 63.9<br>(61.7, 66.1) |

Estimates (95% confidence intervals) are standardized to age distribution in overall sample.
a Among those with self-reported diabetes or high blood glucose ( $\geq 126\text{mg/dL}$ [fasting for 12 hours] or $\geq 220\text{mg/dL}$ [not fasting]).
b Among those with self-reported diabetes ('Diagnosed')

Among those with diagnosed diabetes, the proportion currently taking medication was higher in urban areas (69.4% [67.4, 71.5]) compared to rural areas (52.7%, [51.0, 54.3]). Women were less likely to be receiving treatment than men. The proportion treated was greater with higher age and household wealth, with no consistent differences by attained schooling in both urban and rural areas (**Table 2**).

Among those with diagnosed diabetes, the proportion with controlled diabetes was higher in rural areas (69.4% [68.1, 70.6]) compared to urban areas (59.6% [58.1, 61.0]). Controlled diabetes was higher among women (67.6% [66.4, 68.8]) than men (62.6% [61.4, 63.8]), and adults aged 18-39 years (75.2% [73.6, 76.8]) compared to 45-64 years (60.0% [58.9, 61.1]), but was not greater with higher schooling or household wealth. These socio-demographic disparities were observed in both urban and rural areas (**Supplementary Table 3**). Among those with controlled diabetes, only 47.1% (45.7, 48.4) nationally, 40.5% (38.9, 42.0) in rural areas and 56.8% (54.2, 59.5) in urban areas were taking medication (**Supplementary Table 4**).

### State-level care continuum

Higher diabetes prevalence was observed in urban versus rural areas across all states (**Supplementary Figure 2** and **Supplementary Figure 3**). Diabetes prevalence was higher among the southern states (Kerala, Tamil Nadu, Karnataka, Telangana, Andhra Pradesh), union territories (Andaman & Nicobar Islands, Lakshadweep, Puducherry), and Goa compared to other parts of the country (**Figure 2**; median in rural: 8.8% vs 4.2%; median in urban: 11.4% vs 7.1%). Beyond the regional and state-level heterogeneity observed in **Figure 2**, there were disparities in diagnosis, treatment, and control between socio-demographic groups within each state (interactive dashboard).

**Figure 2.**
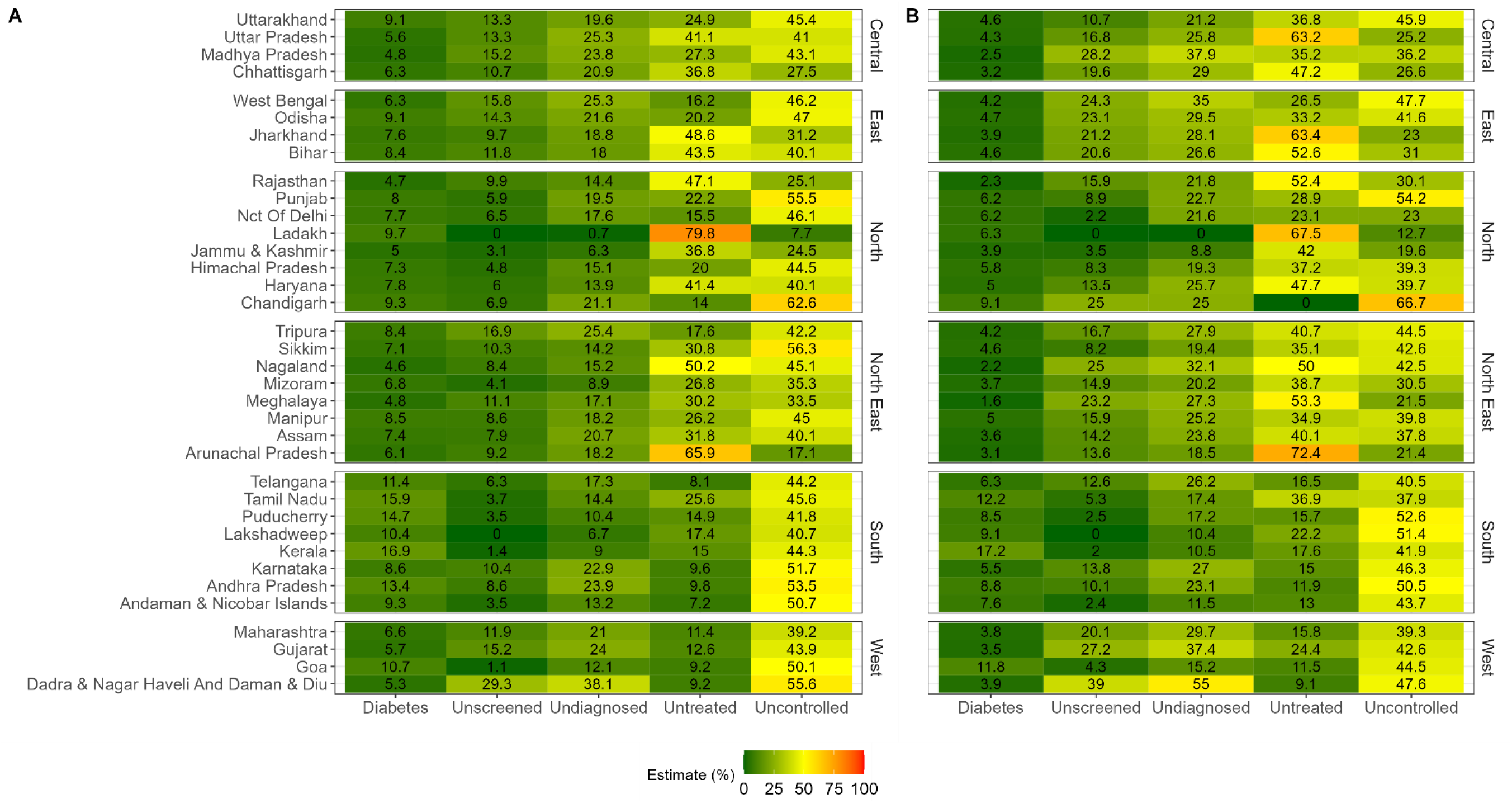
State-level priorities for unmet need in diabetes care cascade, n = 1,651,176. A: Urban, B: Rural; All values are crude percentages. Undiagnosed are among those with diabetes (n = 93,263). Untreated and uncontrolled are among those diagnosed with diabetes (n = 67,209).

Benchmarking state-level diabetes care continuum indicators to the WHO Diabetes Compact targets, the diagnosed diabetes target was met in rural areas of 11 states/UTs, and in urban areas of 23 states/UTs. However, across all states and UTs except Arunachal Pradesh, Ladakh, and Jammu & Kashmir, controlled diabetes was below the 80% target in both urban and rural areas (**Figure 2**).

### District-level care continuum

Between-district variation in the diabetes care continuum were observed in many states. For example, at the district-level, central Kerala (e.g. Kottayam: 22.0 [19.5, 24.6], Ernakulam: 17.8 [15.8, 19.8]) had higher prevalence than north Kerala (e.g. Wayanad: 10.8 [9.4, 12.2], Kasaragod: 11.0 [9.5, 12.5]) although the proportions of those diagnosed (Kottayam: 90.7 [87.5, 93.8], Ernakulam: 94.2 [91.9, 96.5], Wayanad: 90.0 [85.2, 94.7], Kasaragod: 91.0 [87.3, 94.7]) were similar (**Supplementary Figure 4A)**. In Karnataka, among those who were diagnosed, there was substantial between-district heterogeneity in treatment and control. Gulbarga (6.2 [4.3, 8.1]) and Raichur: 4.7 [3.2, 6.2]) had lower prevalence compared to Davanagere (9.2 [7.1, 11.3]), similar levels of low treatment (**Supplementary Figure 4B-C;** Gulbarga: 66.8 [50.4, 83.2], Raichur: 61.9 [45.6, 78.2], Davanagere: 71.2 [56.8, 85.5]) and higher controlled diabetes (Gulbarga: 72.3 [61.8, 82.8], Raichur: 72.7 [61.2, 84.2], Davanagere: 57.9 [44.5, 71.4]). Compared to these districts, Mysuru and Bengaluru had similar prevalence of diabetes (Mysuru: 6.9 [5.7, 8.1], Bengaluru: 9.6 [7.2, 11.9]), higher proportions of treated diabetes (Mysuru: 94.9 [90.8, 99.0], Bengaluru: 90.1 [84.7, 95.5]), and lower proportions of controlled diabetes (Mysuru: 43.2 [29.5, 56.9], Bengaluru: 39.9 [31.0, 48.7]).

There was considerable between-district variation in the diabetes care continuum (**Figure 3**) such that 69.9% of variance in diagnosis, 51.8% of variance in treatment among diagnosed, and 66.8% of variance in control among diagnosed were at the district-level, with the remaining at the state-level.

**Figure 3.**
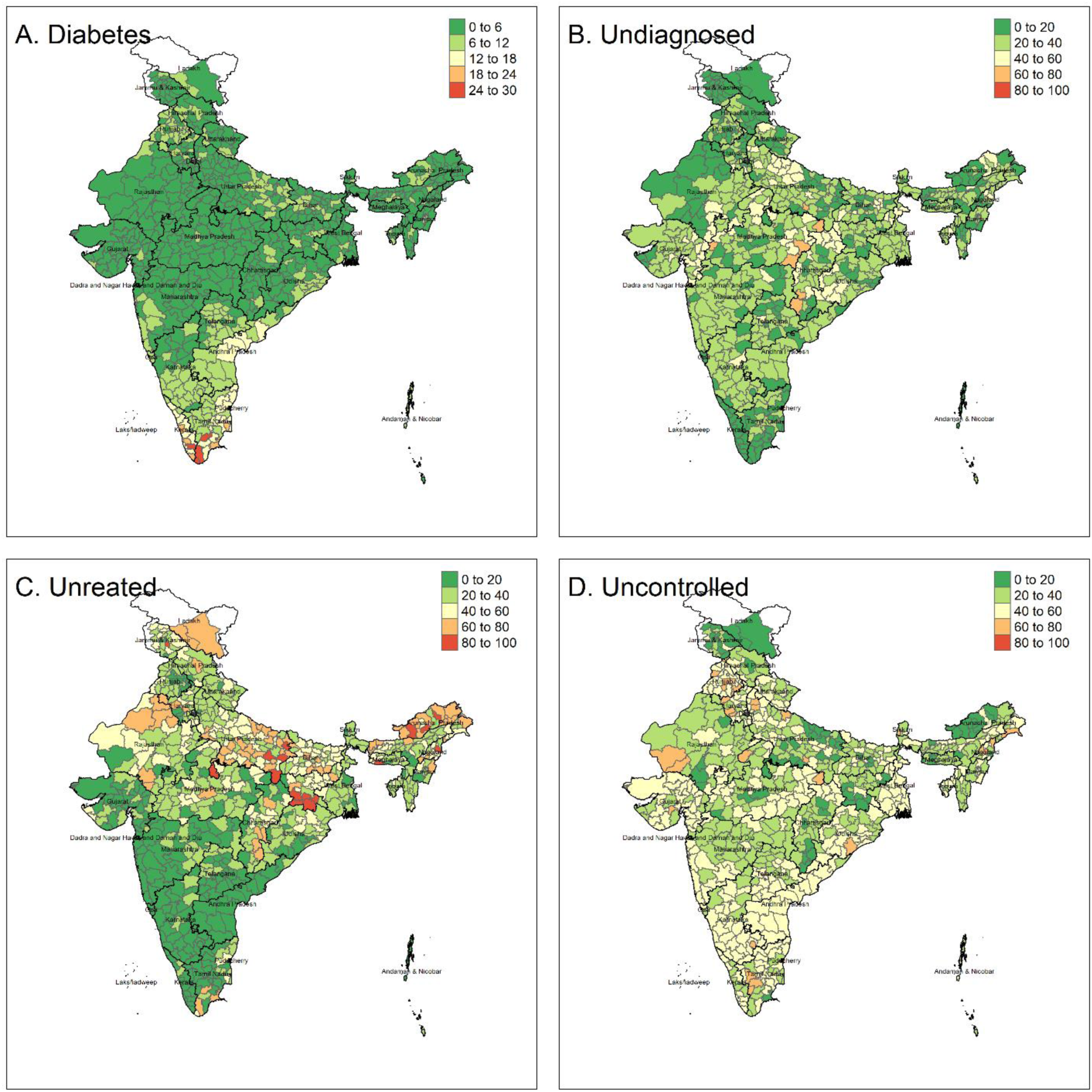
Care cascade in analytic sample by urban and rural residence for 707 districts, n = 1,651,176. All values are crude percentages. Undiagnosed are among those with diabetes (n = 93,263). Untreated and uncontrolled are among those diagnosed with diabetes (n = 67,209). We excluded all districts with less than 50 observations in **Supplementary** Figure 4.

At the district level (n = 707), 246 districts (34.8%) met the 80% diagnosis target while 76 districts (10.7%) met the 80% control target among those diagnosed. Restricting our analysis to districts with at least 50 observations (**Supplementary Figure 5**), consistent with the reporting criteria used in NFHS-5 factsheets, did not change our results (diagnosis target: 36.3%, control target: 10.8%).

## Discussion

Our estimates of the diabetes care continuum in India suggest opportunities for improvement in diagnosis, treatment, and control across all states and districts, and in both rural and urban areas, and across socio-demographic groups. Our analysis suggested three key policy-relevant findings. First, among those with diabetes, nearly 25% were undiagnosed, with lower diagnosis rates in rural areas compared to urban areas. Second, nearly 40% of those with self-reported diabetes were not taking medication, especially in Central, East, North East, and North India – regions where the prevalence is high. Third, 35% of those with diagnosed diabetes did not achieve glycemic control – a finding consistently observed in most states of India. We also observed that the greatest differences in diabetes diagnosis, treatment, and control were between districts in a state, and not between states. The interactive dashboard that accompanies this manuscript highlights these disparities between geographic and socio-demographic subgroups, furthering an agenda of precision public health and identifying district-level priorities for the NMAP for diabetes management. ^7^

The higher proportion of diagnosed and controlled diabetes but lower treated diabetes among women in urban and rural areas suggests greater awareness in urban areas, or potentially better non-pharmaceutical management and maybe lower diabetes severity in rural areas. This may also explain the lower proportions of controlled diabetes among households with higher wealth. The findings on urban-rural differences in diagnosis and control were also consistent with those from the Indian Council of Medical Research – India Diabetes (ICMR-INDIAB) study.^10^ Higher proportions of diagnosed and treated diabetes with higher age, urbanicity, and household wealth were consistent across states and likely associated with greater awareness and access to medical care.

Global studies on diabetes care continuum suggest substantial disparities between- and within-countries. Glycemic control decreased over time from 2005-08 to 2013-16 in USA, with older adults and women more likely to achieve it, compared to younger and middle-aged adults, and men.^14, 26^ An analysis of 28 low and middle income countries reported that 77% of adults experienced an unmet need for diabetes care (undiagnosed, untreated, uncontrolled, and never tested) at some stage of the continuum, with better performance among upper middle income countries.^27^ Nearly half of the outpatients in a retrospective analysis of medical records reported uncontrolled diabetes in eight high income European countries, with considerable between-country variation.^4^

Estimates from this analysis of recent NFHS data show higher proportions of diagnosed diabetes than previous estimates – e.g., among participants aged 18 to 69 years in National Noncommunicable Disease Monitoring Survey (NNMS) 2017-18, 45.8% self-reported being diagnosed. Among those 45 and older in the Longitudinal Aging Study in India (LASI) 2017-18, 60.4% reported being diagnosed.^12, 13^ This may be related to differential classification of diabetes status based on the biomarkers used in these different surveys, greater awareness of diabetes over time, or differential response rates due to the COVID-19 pandemic and sampling strategies – all of these possibilities warrant further analysis that are beyond the scope of this manuscript.^28^ For example, our non-fasted blood glucose cut-off of 220 mg/dL may have high specificity and low sensitivity, identifies only severe diabetes making it more likely to be diagnosed, and therefore, underestimates the true burdens of diabetes and unmet need in diagnosis.

Our estimates for proportions of adults who attained blood glucose control are higher (65.5%) than the ICMR-INDIAB and LASI findings where 36.3% adults older than 20 years and 46.1% adults older than 45 years with self-reported diabetes attained glycemic control (HbA1c <7.0%).^10^ Results from NFHS-4 (2015-16; 15-49 years) and NNMS (2017-18; 18-69 years) also report poor glucose control (NFHS-4: 24.8%, NNMS: 15.7%) among those with diagnosed diabetes.^11, 12^ Beyond possible reasons identified for higher diagnosis, NFHS-5 used random blood glucose to define glycemic control and did not collect HbA1c that is more appropriate for this purpose.

The interactive dashboard permits exploratory analysis to identify those with higher unmet needs among geographic and socio-demographic subgroups. The dashboard also presents a tool for policymakers to prioritize resources across steps in the care cascade for their administrative regions. Similar interfaces ought to be available for monitoring national quality of diabetes care targets set by NMAP and the Global Diabetes Compact.^7, 8^ For example, a state or district health official can navigate to the ‘Socio-demographic Disparities’ and ‘District Disparities’ tabs and compare priorities across different districts within a state, and explore socio-demographic disparities in the care continuum by sex, age, education, and wealth quintile. Additionally, the visualization could help in prioritizing regions for more frequent and widespread screening to improve rates of diagnosis. Investments towards meeting Indian Council of Medical Research’s routine screening guidelines (annual for pre-diabetes, once in 3 years for adults > 30 years) could prove cost effective if it is coupled with higher usage of nearby public healthcare facilities for diabetes management.^29^

This is the most comprehensive report of the diabetes care continuum among adults in a low- and middle-income country at the sub-national level and across socio-demographic subgroups. The most recent data from India (ICMR-INDIAB) reported estimates of glycemic control over 12 years (2008-2020) among adults with self-reported diabetes (n = 5,789 out of 113,043) aged 20 years and older in 30 states and union territories.^9, 10^ Data from the India NNMS 2017-18 also showed national-level care cascades stratified by sex, urbanicity, and age category among adults (18-69 years; n = 9,721).^12^ However, due to its limited sample size, NNMS was not able to present sub-national estimates for socio-demographic groups to appropriately target interventions that could address care gaps. Previous data from the NFHS and LASI were limited to specific age groups and utilized different biochemical markers for glycemia. ^11,13^ Of note, the HbA1c cutoff used (≥ 6.5%) in LASI may overestimate diabetes prevalence among ethnicities other than Non-Hispanic Whites.^28, 30^

Despite its large sample size and representativeness across geographic levels, our analysis has several limitations. First, our analysis is subject to information bias from self-report of high blood glucose and not medical records. However, the prevalence of self-reported diagnosed diabetes in the total population was comparable across different surveys (NFHS-5 vs LASI, INDIAB, and NNMS).^12, 13^ Second, we used a combination of fasting (about 1%) and random blood glucose values to determine diabetes status. Though a single capillary glucose doesn’t meet the confirmatory standards of diagnosis for diabetes (i.e. consecutive elevated glucose levels or elevated glucose and HbA1c levels at the same visit), this approach provides internal consistency and has been used in similar population-based studies.^11, 22^ NFHS-5 also did not collect HbA1c to appropriately define long-term control. Third, the survey was limited in the nature of data it collected for the analytic sample used in this manuscript. For example, BMI and waist circumference data for those older than 49 years in women and 54 years in men prevented us from assessing heterogeneity in care continuum by levels of weight status and whether screening guidelines were met across all ages. We did not have information on duration and family history of diabetes. The survey did not collect data on type of usual care provider and number of visits among those with self-reported diabetes. We were unable to differentiate between types of diabetes. Moreover, we did not have data on older adults living by themselves or institutionalized and non-civilian adults.^18, 20^

India’s rising diabetes burden across all socio-demographic groups present a challenge for public health and healthcare. Initiatives such as NMAP and the NPCDCS offer opportunities for ameliorating this rise through decentralized, targeted interventions. Near real-time monitoring of quality, coverage, and success of these initiatives through performance indicators such as the WHO Diabetes Compact, using interactive dashboards, may be a crucial step in this defeating diabetes.

## Data Availability

All datasets used in this analysis are available for download at www.dhsprogram.com. The code for the analysis is available on https://github.com/jvargh7/nfhs_cascade.

https://egdrc-precision-medicine.shinyapps.io/diabetes_cascade/

## Ethics approval and consent to participate

All participants gave written informed consent before participation. We were exempted from ethical approval for the secondary data analysis from the Institutional Review Board of Emory University.

## Consent for publication

Not applicable

## Author contributions

JSV, PG, NS and MKA developed the study and the analysis plan with inputs from all authors. JSV performed the statistical analysis, prototyped the dashboard and wrote the first draft with inputs from MKA. All authors edited and approved the manuscript

## Acknowledgements

We thank the participants and survey enumerators of National Family Health Survey 2019-21.

---

**Supplementary Figure 1.**
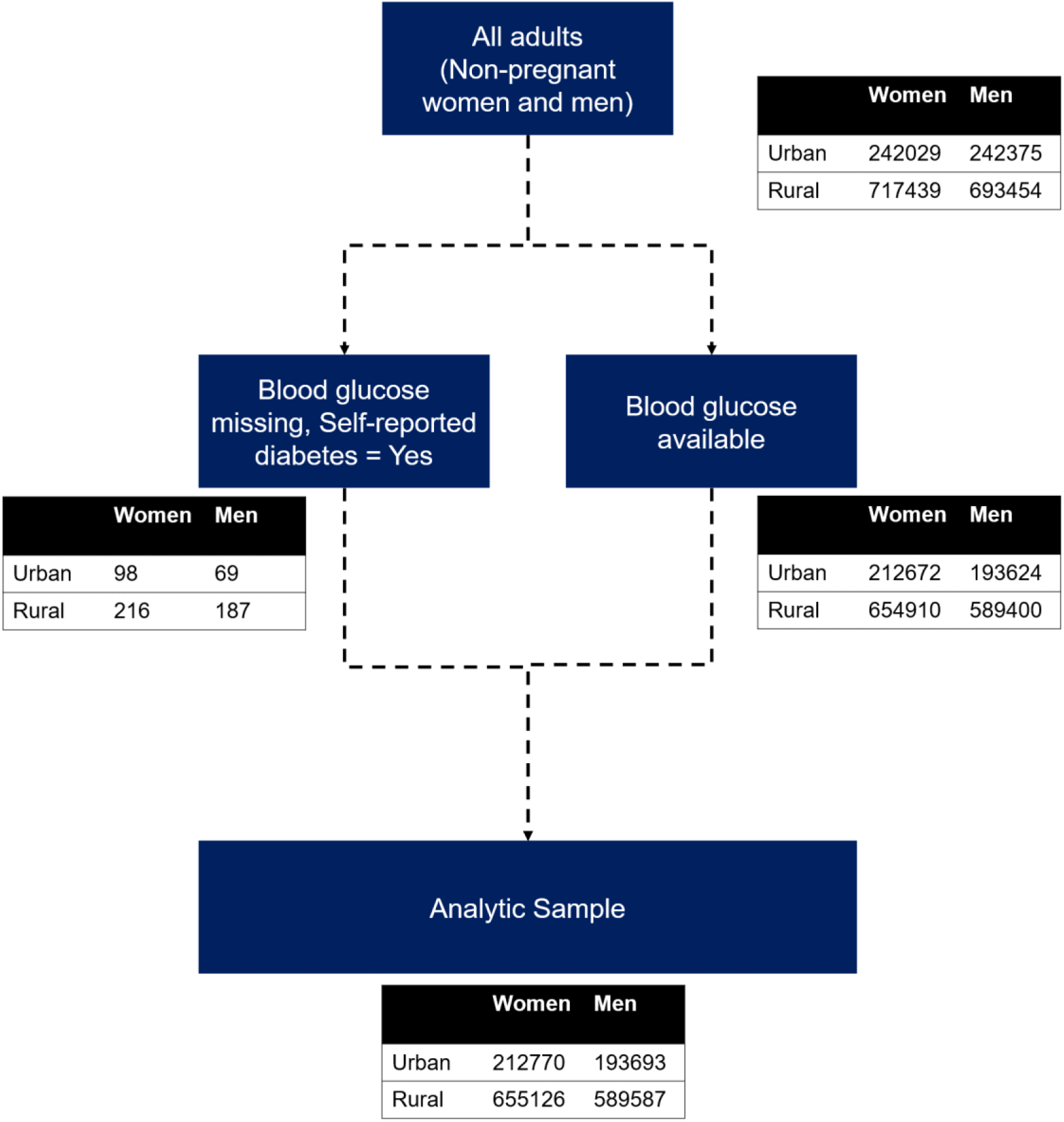
Flowchart of analytic sample.

**Supplementary Figure 2.**
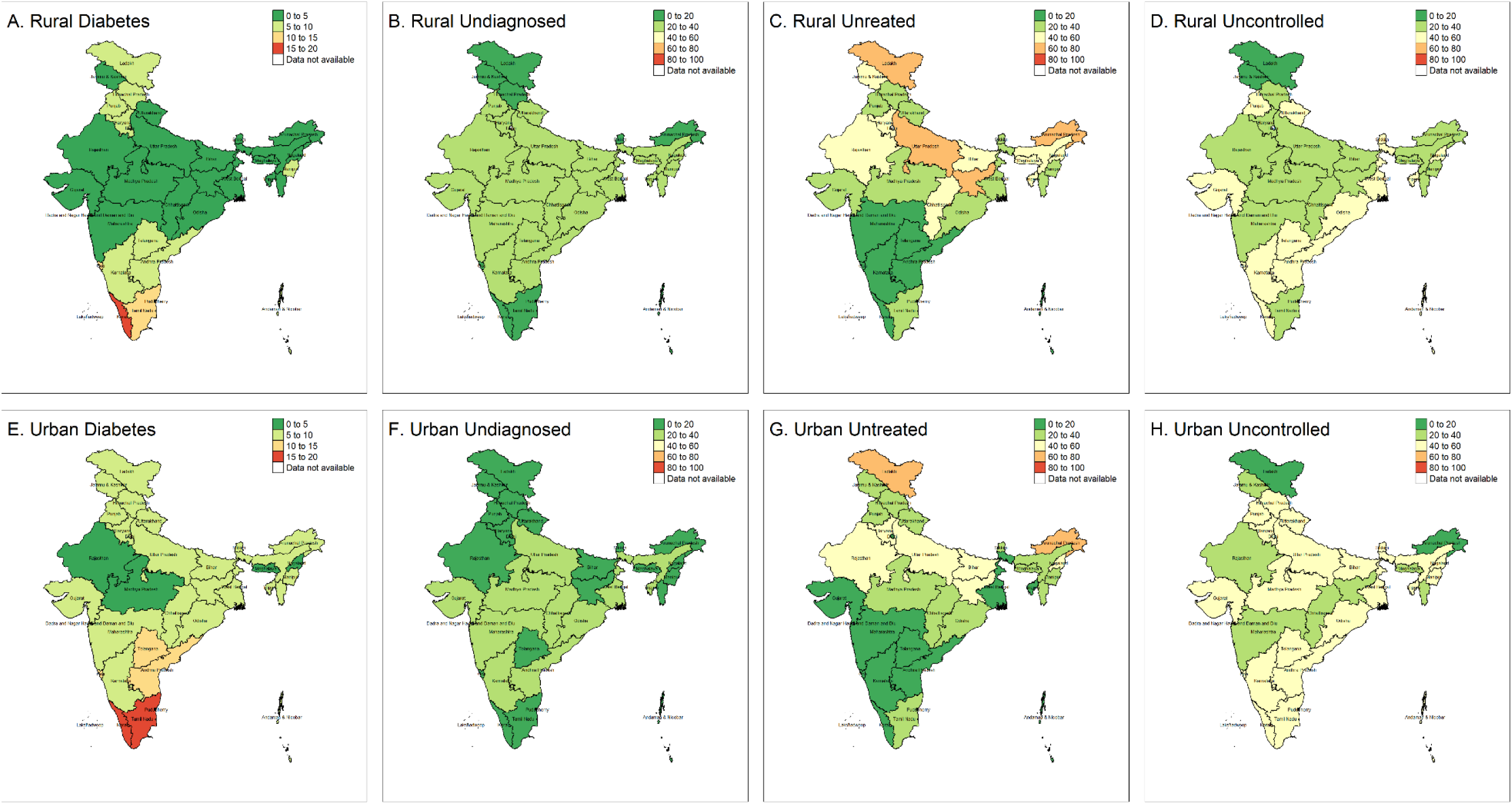
State-level care cascade. Refer to dashboard for detailed presentation; Undiagnosed are among those with diabetes (n = 93,263). Untreated and uncontrolled are among those diagnosed with diabetes (n = 67,209). We excluded all strata with less than 50 participants.

**Supplementary Figure 3.**
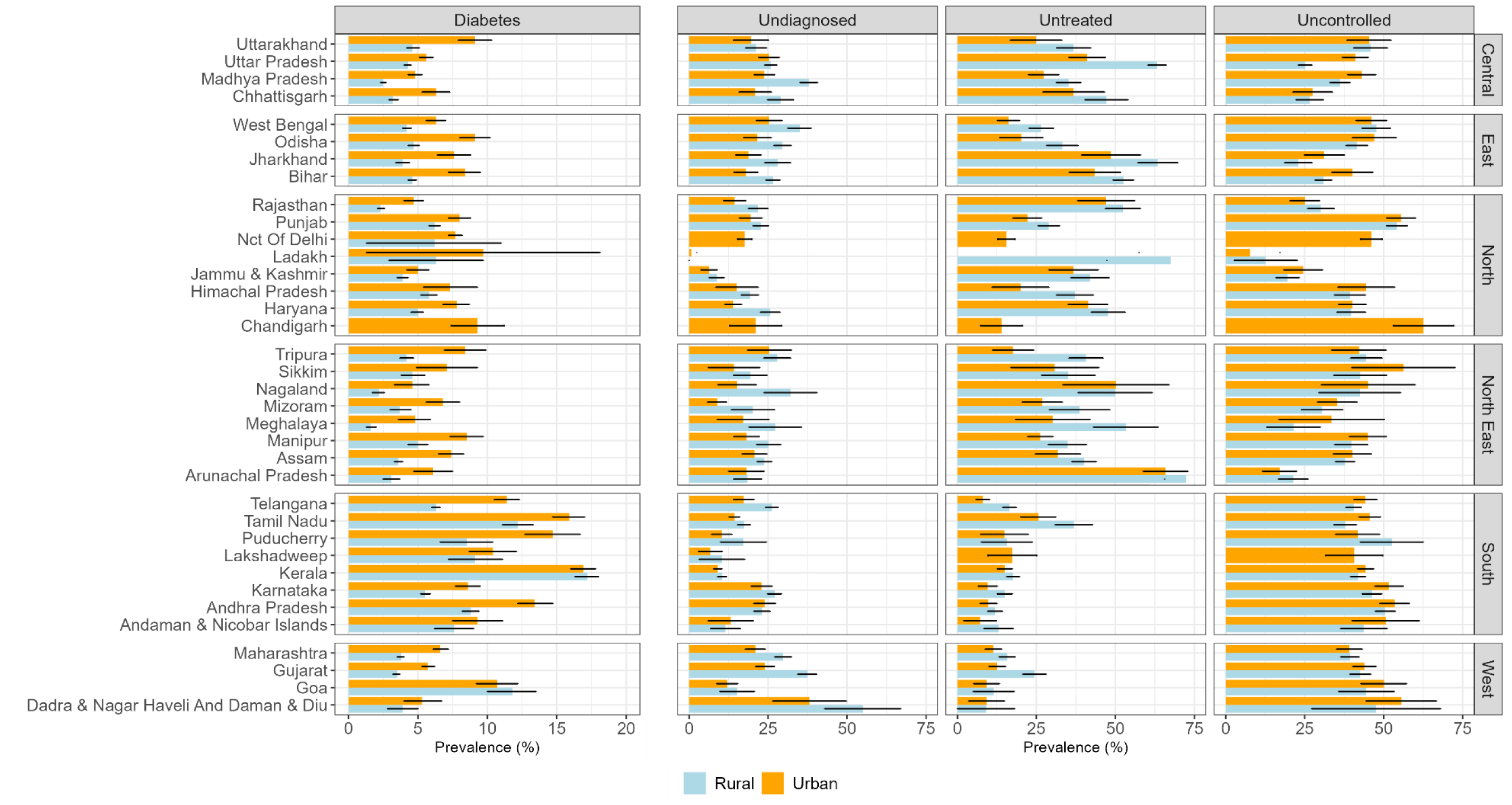
Column plot for care cascade. All values are crude percentages (95% confidence intervals). Undiagnosed are among those with diabetes (n = 93,263). Untreated and uncontrolled are among those diagnosed with diabetes (n = 67,209). We excluded all strata with less than 50 observations.

**Supplementary Figure 4.**
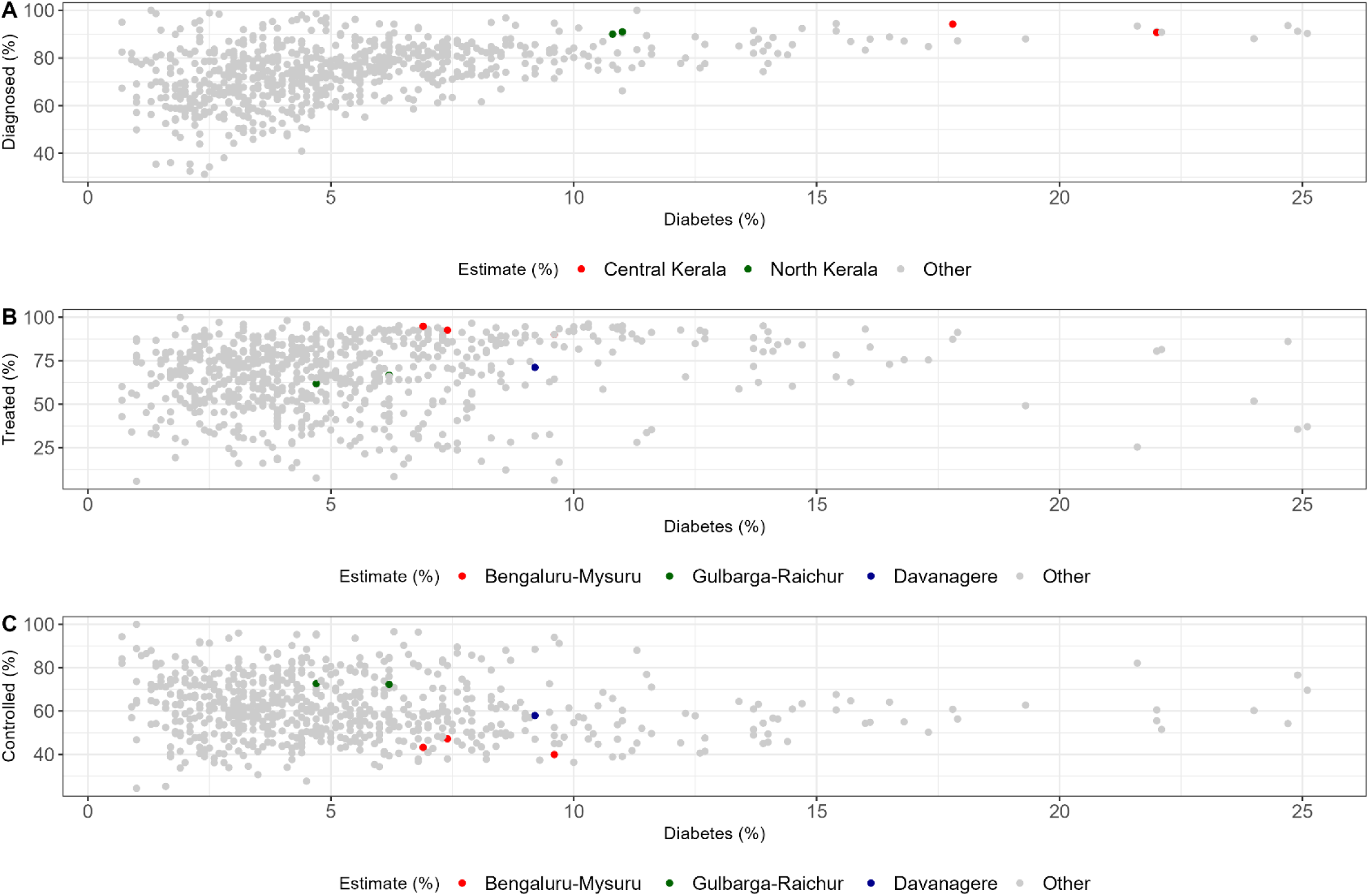
Disparities within states at the district level.

**Supplementary Figure 5.**
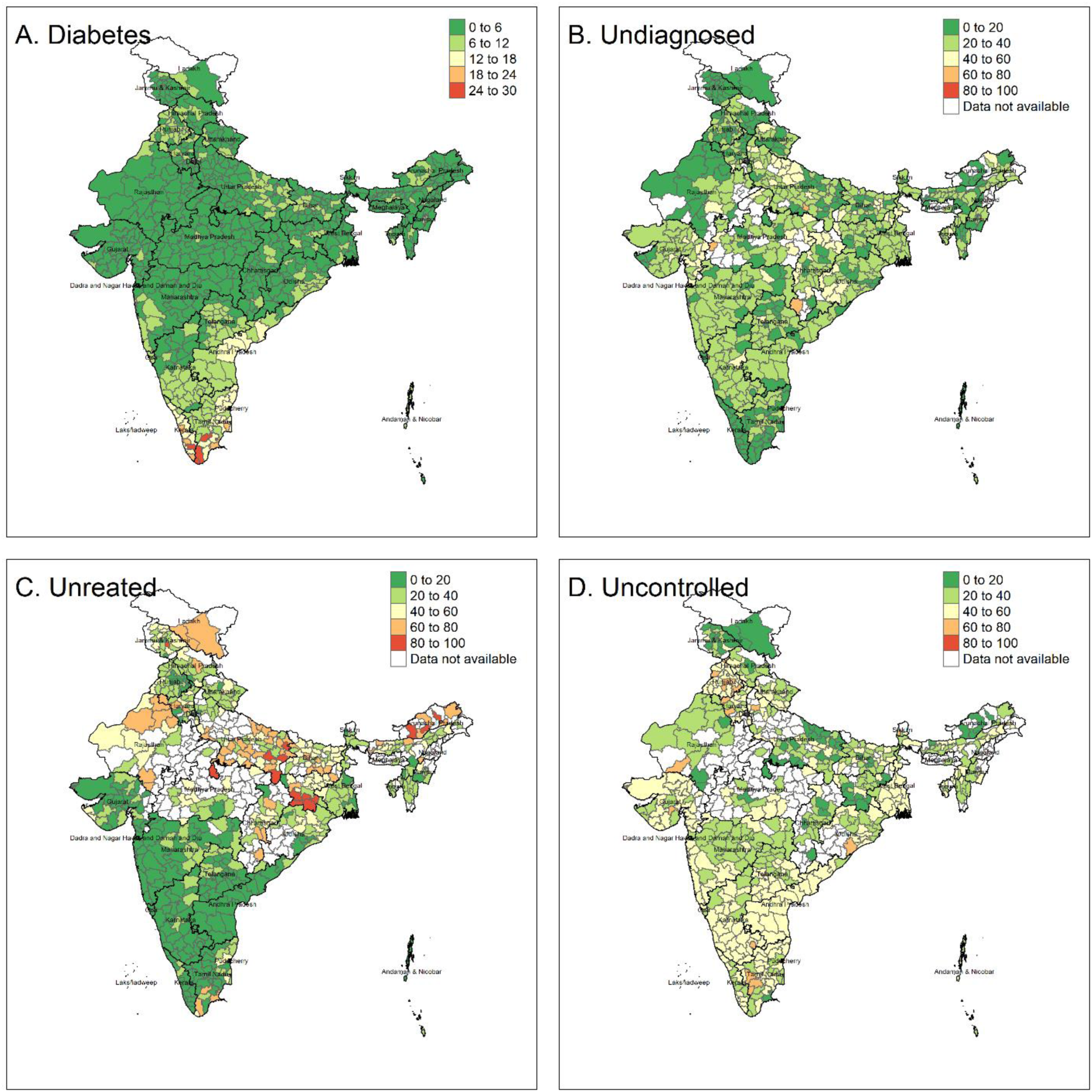
District-level care cascade in analytic sample by urban and rural residence for districts with at least 50 observations. All values are crude percentages.

**Supplementary Table 1.** Definitions of disease and care cascade of diabetes

| Term | Study Population | Definition | Comment |
| --- | --- | --- | --- |
| **Individual-level indicators** |  |  |  |
| Fasting | Analytic sample | (a) Time since last eaten $\geq 12$ hours AND<br>(b) Time since last drank, something other than water $\geq 8$ hours | In our analytic sample, less than 1% are fasted |
| Diabetes | Analytic sample | (a) Self-reported diabetes<br>OR (b) Currently taking medication for diabetes<br>OR (c) High blood glucose ($\geq 126$ mg/dL if fasting or $\geq 220$ mg/dL if not fasting) | A single capillary glucose doesn't meet the confirmatory standards of diagnosis for diabetes (i.e. consecutive elevated glucose levels or elevated glucose and HbA1c levels at the same visit) |
| Screening |  |  | ICMR guidelines recommend annual screening for adults with pre-diabetes and every 3 years for those older than 30 years |
| Diagnosis | "Diabetes" as Yes | Told had high glucose on two or more occasions by a medical provider | Self-reported |
| Treatment | "Diagnosis" as Yes | Currently taking a prescribed medicine to lower glucose | Self-reported. We do not have information on dosage and type of medication used |
| Control | "Diagnosis" as Yes | Blood glucose in non-hyperglycemic range ($< 126$ mg/dL if fasted and $\leq 180$ mg/dL if non-fasted) | We are defining control based on random blood glucose and not HbA1c (%) |
| **Population-level performance indicators** |  |  |  |
| Diagnosis Gap | "Diabetes" as Yes | Percentage Undiagnosed (%) | Meeting WHO Global Diabetes Compact target requires 80% of patients with diabetes to be diagnosed |
| Treatment Gap | "Diagnosis" as Yes | Percentage Untreated (%) |  |
| Control Gap | "Diagnosis" as Yes | Percentage Uncontrolled (%) | Meeting WHO Global Diabetes Compact target requires 80% of patients with diagnosed diabetes to have good control of blood glucose based on HbA1c (%) $< 8.0$ |

**Supplementary Table 2.** Characteristics of participants in analytic sample versus those excluded, n = 1,895,297

|  | **Urban** |  | **Rural** |  |
| --- | --- | --- | --- | --- |
|  | **Analytic Sample**<br>**(n = 406,463)** | **Excluded Sample**<br>**(n = 77,941)** | **Analytic Sample**<br>**(n = 1,244,713)** | **Excluded Sample**<br>**(n = 166,180)** |
| **Sex** |  |  |  |  |
| Women | 52.6 (52.4, 52.8) | 38.5 (37.9, 39) | 53.1 (53, 53.2) | 38.1 (37.8, 38.4) |
| Men | 47.4 (47.2, 47.6) | 61.5 (61, 62.1) | 46.9 (46.8, 47) | 61.9 (61.6, 62.2) |
| **Age category** |  |  |  |  |
| 18-39 | 50.2 (49.8, 50.5) | 52.4 (51.7, 53) | 50.3 (50.1, 50.5) | 52.6 (52.3, 52.9) |
| 40-65 | 39.5 (39.2, 39.7) | 37.7 (37.1, 38.2) | 38.5 (38.3, 38.6) | 35.2 (34.9, 35.5) |
| 65 and above | 10.3 (10.2, 10.5) | 10 (9.6, 10.4) | 11.2 (11.1, 11.3) | 12.2 (12, 12.4) |
| **Schooling** |  |  |  |  |
| None | 16 (15.6, 16.4) | 14.2 (13.6, 14.8) | 33 (32.7, 33.2) | 31.1 (30.7, 31.5) |
| Primary (up to 4<sup>th</sup> class) | 11.9 (11.7, 12.2) | 9.6 (9.2, 10) | 15.5 (15.4, 15.7) | 14 (13.7, 14.2) |
| Secondary (5<sup>th</sup> to 10<sup>th</sup> class) | 47 (46.7, 47.4) | 44.4 (43.6, 45.3) | 41 (40.8, 41.2) | 42.2 (41.8, 42.6) |
| Post-secondary (11<sup>th</sup> class and above) | 25 (24.5, 25.6) | 31.8 (30.6, 32.9) | 10.5 (10.2, 10.7) | 12.7 (12.4, 13) |
| **Caste of head of household** |  |  |  |  |
| General or unspecified | 35.1 (34.2, 36) | 41.5 (39.9, 43.2) | 23.4 (22.9, 23.9) | 24.4 (23.8, 25.1) |
| Other Backward Castes | 41.8 (40.9, 42.7) | 38.6 (37.2, 40) | 41.9 (41.5, 42.4) | 42.6 (41.9, 43.3) |
| Scheduled Caste | 19 (18.3, 19.7) | 16.9 (15.9, 17.9) | 22.7 (22.3, 23.1) | 22.5 (21.9, 23) |
| Scheduled Tribe | 4.1 (3.8, 4.4) | 2.9 (2.6, 3.3) | 12 (11.6, 12.3) | 10.5 (10, 11) |
| **Religion** |  |  |  |  |
| Hindu | 78.7 (77.8, 79.6) | 73.5 (71.9, 75.1) | 84.4 (84, 84.8) | 81.4 (80.7, 82.1) |
| Muslim | 15.4 (14.5, 16.3) | 18.8 (17.2, 20.3) | 10.4 (10, 10.8) | 12.4 (11.8, 13) |
| Other | 6 (5.6, 6.3) | 7.7 (6.8, 8.6) | 5.2 (5, 5.4) | 6.2 (5.8, 6.6) |
| **Household wealth quintile**<br>**(by residence)** |  |  |  |  |
| Lowest | 19.4 (18.7, 20.1) | 13.8 (12.9, 14.8) | 17.8 (17.5, 18.1) | 17.8 (17.3, 18.3) |
| Low | 20.2 (19.7, 20.7) | 16.4 (15.6, 17.2) | 19.1 (18.9, 19.4) | 18.5 (18.1, 18.9) |
| Medium | 20.4 (19.9, 20.8) | 18.2 (17.4, 19) | 20.4 (20.1, 20.6) | 18.8 (18.4, 19.2) |
| High | 20.3 (19.8, 20.8) | 22 (21.1, 23) | 21.2 (20.9, 21.5) | 20.1 (19.7, 20.6) |
| Highest | 19.8 (19.1, 20.5) | 29.6 (27.9, 31.2) | 21.5 (21.1, 21.8) | 24.7 (24, 25.4) |
| **WHO body mass index categories**<br>**(18-49y in women, 18-54y in men)** |  |  |  |  |
| Underweight (<18.5 kg/m<sup>2</sup>) | 4.4 (4.2, 4.5) | 0.3 (0.3, 0.4) | 7.4 (7.3, 7.5) | 0.7 (0.6, 0.7) |
| Normal (18.5-24.9 kg/m<sup>2</sup>) | 22.7 (22.5, 23) | 1.8 (1.6, 2) | 25.4 (25.3, 25.6) | 2.4 (2.2, 2.5) |
| Overweight (25-29.9 kg/m<sup>2</sup>) | 10.2 (10, 10.4) | 1.2 (1.1, 1.3) | 6.9 (6.9, 7) | 1.1 (1, 1.1) |
| Obese (≥30 kg/m<sup>2</sup>) | 4.3 (4.2, 4.5) | 0.6 (0.5, 0.7) | 1.9 (1.9, 2) | 0.4 (0.3, 0.4) |
| **High waist circumference**<br>**(18-49y in women, 18-54y in men)** | 49.1 (48.4, 49.8) | 60.7 (58.3, 63.2) | 35.5 (35.2, 35.8) | 46 (44.4, 47.6) |
| **Diabetes Care Cascade** |  |  |  |  |
| Fasting (%) | 1.2 (1.2, 1.3) |  | 1.3 (1.3, 1.4) |  |
| Diabetes in total population (%) | 8.4 (8.2, 8.6) |  | 5 (4.9, 5.1) |  |
| Diagnosed in total population (%) | 6.8 (6.6, 7) |  | 3.8 (3.7, 3.9) |  |
| Treated in total population (%) | 5.3 (5.2, 5.5) |  | 2.5 (2.4, 2.5) |  |
| Controlled in total population (%) | 3.8 (3.7, 4) |  | 2.4 (2.3, 2.4) |  |

All values are percentages (95% confidence intervals). Height and weight were measured using SECA 213 Stadiometer and SECA 874U digital scale respectively for men 15-54 years eligible for the interview. Body mass index was categorized based on WHO cut-offs (underweight: <18.5 kg/m<sup>2</sup>, overweight: ≥ 25-29.9 kg/m<sup>2</sup>, obesity: ≥ 30kg/m<sup>2</sup>) (WHO Tech Rep Ser 1995). Gulick tape was used to measure waist circumference for men 15-54 years eligible for the interview. High waist circumference (yes or no) was categorized based on WHO sex-specific cut-offs (men: ≥94 cm, women: ≥80 cm) (Misra 2006 Int J Obes).

**Supplementary Table 3.** Care cascade estimates conditional on previous step in India, 2019-21

|  | Urban |  |  |  | Rural |  |  |  |
| --- | --- | --- | --- | --- | --- | --- | --- | --- |
|  | Diabetes (%) | Diagnosed <sup>a</sup> (%) | Treated <sup>b</sup> (%) | Controlled <sup>c</sup> (%) | Diabetes (%) | Diagnosed <sup>a</sup> (%) | Treated <sup>b</sup> (%) | Controlled <sup>b</sup> (%) |
| **Total** | 9.7<br>(9.4, 9.9) | 77.2<br>(76, 78.4) | 69.4<br>(67.4, 71.5) | 50.4<br>(48.4, 52.3) | 4.9<br>(4.8, 5) | 72.2<br>(71.1, 73.3) | 52.7<br>(51, 54.3) | 52.9<br>(51.4, 54.5) |
| **Sex** |  |  |  |  |  |  |  |  |
| Women | 9.1<br>(8.8, 9.4) | 78.6<br>(77, 80.2) | 68<br>(65.6, 70.5) | 50.8<br>(48.1, 53.4) | 4.3<br>(4.2, 4.4) | 74.2<br>(72.7, 75.6) | 50<br>(47.9, 52) | 53<br>(51, 55) |
| Men | 10.2<br>(9.9, 10.6) | 75.6<br>(74.1, 77.1) | 71.2<br>(69, 73.3) | 49.9<br>(47.2, 52.6) | 5.6<br>(5.5, 5.8) | 69.7<br>(68.3, 71.2) | 56.4<br>(54.6, 58.3) | 52.9<br>(50.7, 55.1) |
| **Age category** |  |  |  |  |  |  |  |  |
| 18-39 | 2.4<br>(2.3, 2.6) | 70.5<br>(67.9, 73) | 51.2<br>(47.6, 54.8) | 53.7<br>(49.9, 57.6) | 1.8<br>(1.7, 1.8) | 69.7<br>(67.9, 71.5) | 39.9<br>(37.5, 42.2) | 57.8<br>(55.1, 60.5) |
| 40-65 | 12.6<br>(12.3, 13) | 80.3<br>(79.4, 81.2) | 80.3<br>(78.9, 81.7) | 45.2<br>(43.8, 46.5) | 7.8<br>(7.6, 7.9) | 74.1<br>(73.3, 74.8) | 68.6<br>(67.4, 69.9) | 45.5<br>(44.4, 46.6) |
| 65 and above | 22.9<br>(22, 23.8) | 87.4<br>(86.3, 88.5) | 85.4<br>(84, 86.8) | 54.7<br>(52.9, 56.6) | 11.9<br>(11.6, 12.3) | 80.3<br>(79.4, 81.3) | 73.6<br>(72.2, 75) | 54.8<br>(53.4, 56.3) |
| **Schooling** |  |  |  |  |  |  |  |  |
| None | 7.4<br>(7, 7.7) | 72.8<br>(69.7, 76) | 68.1<br>(63.7, 72.4) | 47.3<br>(43.8, 50.8) | 3.9<br>(3.8, 4) | 68.9<br>(66.7, 71.1) | 54.4<br>(51.6, 57.2) | 52.7<br>(50.5, 54.9) |
| Primary (up to 4<sup>th</sup> class) | 9.8<br>(9.4, 10.3) | 75.1<br>(71.6, 78.6) | 70.1<br>(66.2, 74) | 47.3<br>(43.6, 51.1) | 5<br>(4.8, 5.2) | 71.6<br>(69.3, 74) | 54.1<br>(50.8, 57.4) | 51.2<br>(47.9, 54.5) |
| Secondary (5<sup>th</sup> to 10<sup>th</sup> class) | 10.1<br>(9.8, 10.5) | 78.1<br>(76.7, 79.5) | 70.7<br>(68.5, 72.9) | 48.5<br>(45.8, 51.3) | 5.5<br>(5.3, 5.6) | 74<br>(72.8, 75.3) | 52.5<br>(50.6, 54.3) | 52.1<br>(49.8, 54.5) |
| Post-secondary (11<sup>th</sup> class and above) | 10<br>(9.5, 10.5) | 80.9<br>(79.1, 82.6) | 67.1<br>(64.3, 69.9) | 56.8<br>(52.3, 61.3) | 6.3<br>(6, 6.6) | 78<br>(75.9, 80.2) | 44.5<br>(41.5, 47.4) | 58.3<br>(53.4, 63.2) |
| **Household wealth quintile** |  |  |  |  |  |  |  |  |
| Lowest | 6.3 | 69.7 | 57.5 | 54.2 | 2.8 | 65.8 | 41.4 | 65.7 |
|  | (5.9, 6.7) | (66.5, 72.9) | (52.7, 62.3) | (49.4, 59) | (2.6, 2.9) | (63.1, 68.4) | (38, 44.8) | (61.4, 70) |
| Low | 8.9<br>(8.5, 9.3) | 72.2<br>(69.8, 74.7) | 67.4<br>(64.1, 70.6) | 48.6<br>(44.7, 52.6) | 3.5<br>(3.3, 3.6) | 69.4<br>(67.1, 71.8) | 45.7<br>(42.5, 48.8) | 59.4<br>(55.5, 63.3) |
| Medium | 10.2<br>(9.7, 10.7) | 78.2<br>(76.2, 80.1) | 72.5<br>(69.6, 75.4) | 47.3<br>(43.3, 51.4) | 4.5<br>(4.3, 4.6) | 72<br>(70, 74.1) | 47.5<br>(44.8, 50.3) | 54.8<br>(51.5, 58.2) |
| High | 11.1<br>(10.6, 11.6) | 80.1<br>(77.8, 82.4) | 70.8<br>(67.7, 74) | 51.6<br>(47.9, 55.3) | 5.8<br>(5.7, 6) | 71.4<br>(69.2, 73.6) | 55.6<br>(52.9, 58.4) | 52.4<br>(49.6, 55.1) |
| Highest | 11.6<br>(11.1, 12.1) | 81.6<br>(79.2, 84) | 72.4<br>(69.2, 75.6) | 51.3<br>(47.1, 55.5) | 7.6<br>(7.4, 7.8) | 75.7<br>(73.8, 77.6) | 58.2<br>(55.6, 60.8) | 49<br>(46.3, 51.6) |

Estimates (95% confidence intervals) are standardized to age distribution in overall sample.
a Among those with self-reported diabetes or high blood glucose ( $\geq 126$ mg/dL [fasting for 12 hours] or $\geq 220$ mg/dL [not fasting]).
b Among those with self-reported diabetes
c Among those seeking treatment with self-reported diabetes (Urban: 19423, Rural: 25580) defined using blood glucose ( $< 126$ mg/dL [fasting for 8 hours] or $\leq 180$ mg/dL [not fasting])

**Supplementary Table 4.** Proportion of those diagnosed and treated among those with controlled blood glucose

|  | **Total** | **Urban** | **Rural** |
| --- | --- | --- | --- |
|  | **Treated (%)** | **Treated (%)** | **Treated (%)** |
| **Total** | 47.1 (45.7, 48.4) | 56.8 (54.2, 59.5) | 40.5 (38.9, 42) |
| **Sex** |  |  |  |
| Women | 45.2 (43.6, 46.7) | 55.9 (52.8, 58.9) | 37.9 (36.1, 39.7) |
| Men | 49.5 (47.9, 51) | 58.1 (55.2, 61) | 43.7 (41.8, 45.6) |
| **Age category** |  |  |  |
| 18-39 | 30.4 (28.5, 32.2) | 37.7 (33.9, 41.6) | 26.3 (24.4, 28.2) |
| 40-65 | 61.8 (60.5, 63.2) | 70.2 (68.1, 72.4) | 55.1 (53.4, 56.8) |
| 65 and above | 71.2 (69.8, 72.6) | 80 (78, 82) | 63.9 (62, 65.8) |
| **Schooling** |  |  |  |
| None | 51.9 (50.1, 53.6) | 62.9 (59.4, 66.3) | 47.8 (45.8, 49.9) |
| Primary (up to 4<sup>th</sup> class) | 53.7 (51.5, 55.8) | 65.2 (61.2, 69.2) | 47.2 (44.5, 49.9) |
| Secondary (5<sup>th</sup> to 10<sup>th</sup> class) | 46.3 (44.6, 47.9) | 57.4 (54.3, 60.5) | 38.6 (36.6, 40.5) |
| Post-secondary (11<sup>th</sup> class and above) | 39.9 (37.2, 42.5) | 49.5 (45.4, 53.6) | 28.2 (25.1, 31.3) |
| **Household wealth quintile** |  |  |  |
| Lowest | 39.4 (36.4, 42.4) | 46 (40.7, 51.4) | 33 (29.9, 36.2) |
| Low | 43.5 (41.2, 45.8) | 53 (48.8, 57.2) | 35.4 (32.6, 38.1) |
| Medium | 46.2 (44, 48.4) | 58.5 (54.4, 62.6) | 37.3 (34.7, 39.8) |
| High | 49.4 (47.4, 51.5) | 59.8 (56, 63.6) | 42.6 (40.1, 45.1) |
| Highest | 50.2 (48.2, 52.3) | 61 (56.9, 65.2) | 44.8 (42.4, 47.2) |

